# The Contribution of Age Structure to the Number of Deaths from Covid-19 in the UK by Geographical Units

**DOI:** 10.1101/2020.04.16.20067991

**Authors:** Hill Kulu, Peter Dorey

## Abstract

This study investigates the contribution of population age structure to mortality from Covid-19 in the UK by geographical units. We project death rates at various spatial scales by applying data on age-specific fatality rates to the area’s population by age and sex. Our analysis shows a significant variation in the projected death rates between the constituent countries of the UK, between its regions and within regions. First, Scotland and Wales have higher projected fatality levels from Covid-19 than England, whereas Northern Ireland has lower rate. Second, the infection fatality rates are projected to be substantially higher in small towns and rural areas than those in large urban areas. Third, our analysis shows that within urban regions there are also ‘pockets’ of high projected death rates. Overall, the areas with high and low fatality rates tend to cluster because of the high residential separation of different population age-groups in the UK. Our analysis also reveals that the Welsh-, Gaelic- and Cornish-speaking communities with relatively old populations are likely to experience heavy population losses if the virus spreads widely across the UK.

## Background

The rapid spread of Covid-19 has become a major public health threat and challenge for many countries. The most recent data show that there are 2.1 million reported cases and more than 139 thousand deaths in the World (1). Studies show that hospital admissions and deaths from Covid-19 significantly vary across population subgroups; severe illness and deaths are more likely to occur among those with underlying health conditions including respiratory diseases such as chronic obstructive pulmonary disease, asthma, but also cardiovascular disease and diabetes (2). Unsurprisingly, the hospital admissions and deaths from Covid-19 exhibit a distinct age pattern (3). The infection fatality rates (IFR) are very low among young, they increase with age and are the highest among those in their eighties. The estimates of the IFR significantly vary across studies; some studies show that as many as a fifth of the oldest population die if infected, other studies report lower levels (4,5). The main issue has been the lack of information on the risk population (or the denominator); the virus testing levels vary by countries, further while there are some estimates on symptomatic cases, the number of asymptomatic cases, i.e. individuals who carry the virus, but exhibit no symptoms are not detected and reported (5).

Countries with older populations are likely to observe higher hospitalisation and mortality rates than those with young populations during the Covid-19 pandemic (6). Although many factors may influence mortality levels from Covid-19 such as well-functioning healthcare system and various prevention measures (e.g. social distancing), a recent study has shown that more than a half of the variation in mortality rates across countries comes from the differences in the population age structure (7). Population age structure varies across countries, but even more within countries across regions. Hence, with the same infection rate mortality levels are expected to be much higher in regions with old populations (e.g. rural areas) than in areas where young populations dominate (e.g. urban areas); further significant differences in the population age structure may also exist within (e.g. urban) regions. A recent study from the UK shows a substantial spatial polarisation between young and old age groups (8). Further the residential separation increases with age, with the highest spatial separation found among those aged 80 and older. This is not surprising as the age is an important determinant of residential choice and location and the concentration of the oldest groups in retirement and nursing homes and communities are well-known pattern in high-income countries. Interestingly, however, residential separation between older and younger generations has risen rapidly in the past three decades in the UK with emerging clusters of the oldest-old population (i.e. aged 85 and older) (8). Although the spatial segregation between the young and the old may limit the spread of the virus within the UK, it is simultaneously clear that it will certainly make some communities much more vulnerable to Covid-19 pandemic than others.

The objective of this study is to investigate how mortality from Covid-19 is expected to vary across spatial units in the UK, to determine spatial clustering of the projected deaths from Covid-19 and to identify vulnerable communities. We extend previous research on the effect of age structure across countries and different regions in Europe (6,9). To our best knowledge, this is the first study to investigate the possible effect of population age structure on mortality from Covid-19 within a country between and within regions. This includes the analysis of patterns across the smallest spatial units to identify spatial clusters and vulnerable communities.

## Method

We calculate the expected number of deaths by geographical units by applying the age-specific fatality rates to the risk population. 

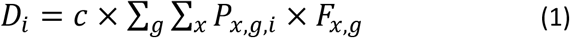

where *D*_*i*_ is the number of expected deaths in geographical unit *i, P*_*x,g,i*_ is the number of individuals aged *x* in sex *g* in a geographical unit, *F*_*x,g*_ is the infection fatality rate (IFR) in age *x* by sex, which is the same for all spatial units, and *c* is the infection rate. We apply the same age-specific fatality rates by sex to all geographical units in the country. We use the IFRs obtained from the study by Ferguson et al. on the UK, which, in turn, are based on the analysis of Covid-19 mortality in China by Verity et al (5,10). We calculate age-specific fatality rates by sex using the latest data on observed deaths by sex in Great Britain (11)^1^. Although Ferguson et al. show lower overall IFR (i.e. 0.9%) than that estimated by many other studies including the estimates by the WHO, they have been adjusted to under-ascertainment (as much as this is possible). Many studies divide the deaths from Covid-19 to reported cases, which significantly over-estimate the IFR. We assume a uniform infection rate, and use the values of 0.2, 0.4 and 0.6 in our calculations for total deaths. Although there are significant differences in the infection rate across geographical units, especially in the beginning of the spread of infection diseases, we assume that many people will be infected by the virus in the long-term, either during the first wave or possible subsequent waves, which are likely to follow within a year or two. The idea is consistent with the findings of the studies on the Spanish flu from 1918 to 1920, which have shown that many countries experienced two or even more waves of the virus before the majority of a population became immune (12–14). Further our main task is to identify areas where people are at high risk because of their characteristics (i.e. old age) and to inform policy-makers with the aim to find the ways of reducing transmission of the virus to these communities.

Next, we calculate the number of deaths per 1,000 population for each geographical unit and thereafter the infection fatality rate relative to that of a reference population, which is a country’s population. 

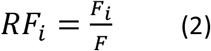

where *RF*_*i*_ is the relative infection fatality rate (or the rate ratio) for geographical unit *i, F*_*i*_ is the fatality rate (per 1,000) for a geographical unit and *F* is the fatality rate for the country.

We use Moran’s *I* statistics to measure the spatial clustering of infection fatality rates. Moran’s *I* is calculated as follows (15,16): 

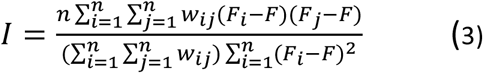

where *n* is the number of spatial units, *F*_*i*_ and *F*_*j*_ are log fatality rates for geographical units *i* and *j, F* is the country’s log fatality rate and *w*_*ij*_ is a measure of the spatial proximity between spatial units *i* and *j*. We use a binary connectivity definition where *w*_*ij*_ = 1 if spatial units *i* and *j* share a common boundary, and *w*_*ij*_ = 0 otherwise. The interpretation of Moran’s *I* is somewhat similar to the Pearson’s correlation coefficient. The value of 1 shows the perfect spatial clustering of similar values (or separation of different values, e.g. mortality rates above the average are all in the North of a country and rates below the average are in the South), whereas the value of 0 shows no spatial autocorrelation in the variable of interest.

Moran’s *I* provides a single summary value of the overall spatial clustering of the area data. However, the spatial clustering itself may vary across space. To assess whether clustering occurs around particular locations and to identify these locations we can use local measures of spatial autocorrelation. We use the local Moran’s statistic to detect spatial clustering of similar values: 

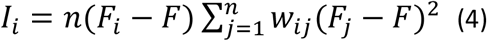

where *I*_*i*_ is the value of Moran’s statistics for geographical unit *i*. The global Moran’s *I* is the sum of the local Moran’s statistics, i.e. *∑ I*_*i*_ *= I* (15,16). We adjust the significance levels by applying the Bonferroni correction, which is an adjustment made to *p*-values when several statistical tests are performed simultaneously on the same data-set (17).

## Data

We use mid-year population by age and sex (one-year intervals) in England, Wales and Scotland by local authority or council areas. Our data come from 2018; this is the latest year we have a detailed information on population age-sex structure by geographical units (18,19). We then repeat our calculations using mid-year population by age and sex for the census lower super output areas (LSOA). The lower output areas are the smallest spatial units for which data are available in the UK; most of them have about 1,000 people. We also use the data on LSOAs to calculate the IFR by area type. We use the urban-rural classification provided by the Office for National Statistics (England and Wales) and National Records of Scotland (20,21). All analyses were performed using R (22–27).

## Results

### Country level

We have calculated the number of expected deaths in the UK and its constituent countries: England, Wales, Scotland and Northern Ireland using age-specific infection fatality rates by sex and different infection prevalence rates. With an infection rate of 0.6 the UK is expected to observe 481 thousand deaths. This figure is very large, but within the same magnitude as the estimates based on a microsimulation model by Ferguson et al., which predicts 510 thousand deaths in Great Britain (for the ‘do-nothing’ scenario) (10). With infection rates of 0.4 and 0.2 the projected number of deaths in the UK are 320 and 160 thousands, accordingly. Although the UK has witnessed a significant increase in the number of deaths from Covid-19 in the recent two weeks, our projected figures still look very large in comparison to the current number of deaths, which is 13,729 (16/04/20) (1). This suggest that either the virus is still much less spread in the total population than believed (28) or that even our conservative (i.e. relatively low) infection fatality rates substantially overestimate the number of deaths. Therefore, in what follows next, we will report the relative distribution of deaths and relative fatality rates in our comparisons as these are immune to the total number of deaths (unless the infection fatality rates by age change significantly).

We have calculated the number of individuals requiring hospitalisation, critical care and the number of deaths by age and sex. As the hospital admission, critical care and fatality rates follow a distinct age pattern, the projected number of hospitalisations and deaths are expected to occur much more in certain age groups than in others (10). Figure 1 shows that there are a relatively few individuals who need hospitalisation among population aged below 30. Their number increases significantly by age and those in their seventies form the largest group of individuals requiring hospitalisation. The distribution of individuals requiring critical care and deaths are even more shifted toward older age groups. Very few individuals younger than age 50 need critical care or are expected to die from Covid-19; most critical care cases and deaths occur among those in their seventies, eighties and nineties – these form 72% of the critical care cases and 71% of the total deaths.

**Figure 1.**
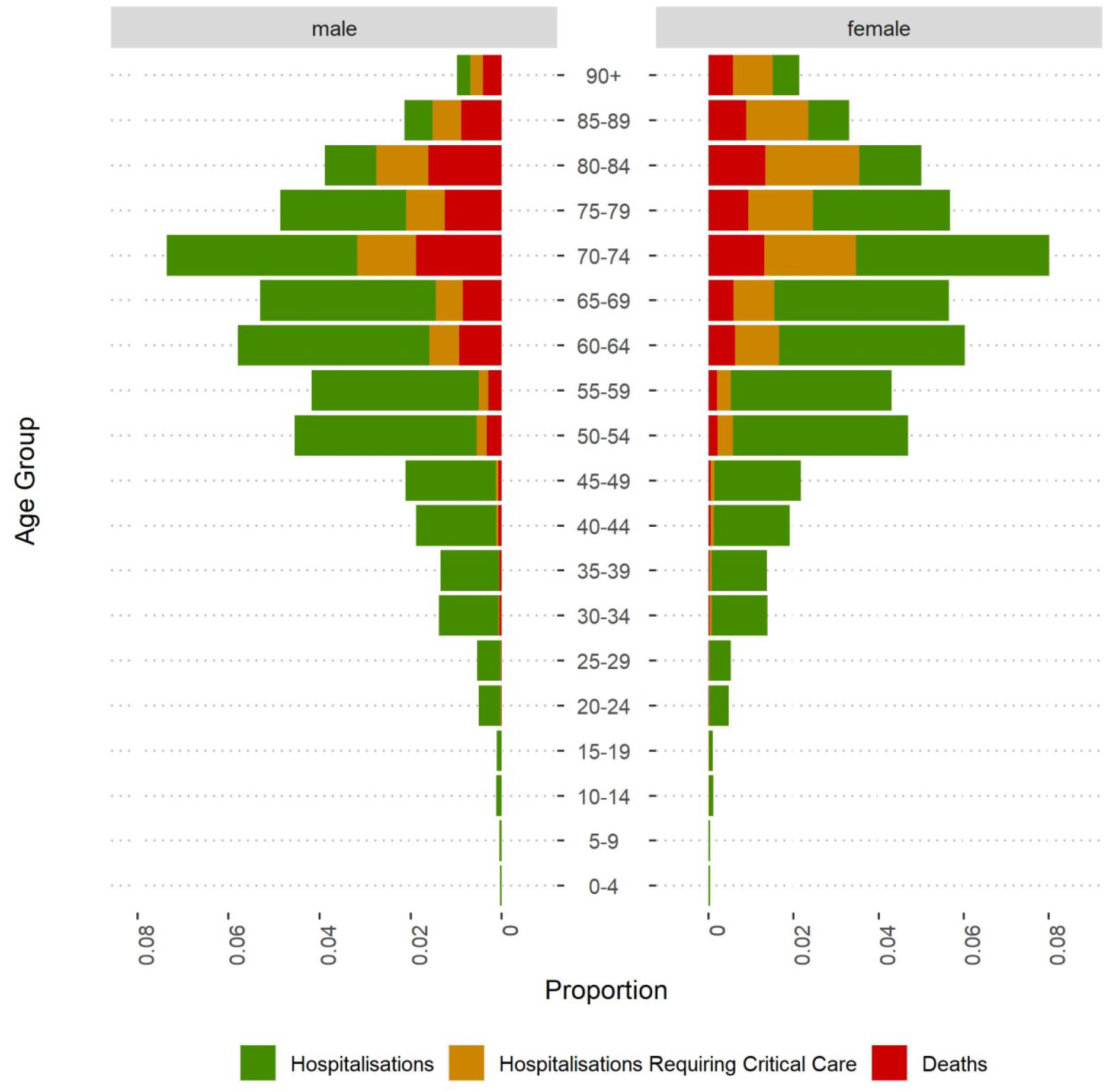
Projected distribution of hospitalised individuals, those requiring critical care and deaths from Covid-19 in Great Britain by age and sex.

Table 1 reports relative hospitalisation, critical care and death rates by the UK’s constituent countries. We see that Scotland and Wales have higher projected hospitalisation rates than England (the comparison group), whereas the rates are substantially lower in Northern Ireland. Largely similar picture emerges when we compare the expected number of deaths from Covid-19. Scotland exhibits 3% and Wales as much as 12% higher fatality rate than England; Northern Ireland has 9% lower rate. These differences reflect significant differences in the population age structure across the UK’s countries. Clearly, Northern Ireland has the youngest age structure, whereas Scotland and Wales have the oldest. The current population age-structures reflect the past demographic processes in each of the four countries. Until the turn of the century, Scotland experienced significant out-migration of young adults; fertility levels have been lower than in England for decades and the country has received fewer international migrants (29,30). Significant out-migration and low immigration also explain a relatively old population structure in Wales. In contrast, Northern Ireland has long experienced substantially higher fertility levels that the rest of the UK and thus has a markedly younger population.

**Table 1.** Projected Relative Hospitalisation and Infection Fatality Rates from Covid-19 in the UK.

| Country | Hospitalisation | Critical care | Fatality |
| --- | --- | --- | --- |
| England | 1 | 1 | 1 |
| Wales | 1.08 | 1.12 | 1.12 |
| Scotland | 1.04 | 1.03 | 1.03 |
| Northern Ireland | 0.94 | 0.91 | 0.91 |

Notes: Relative to England's rates, which are 15.2, 5.0 and 2.4 per 1,000 with IR=0.2

### Local authority level

Next, we have calculated relative infection fatality rates by local authorities in Great Britain (England, Wales and Scotland). As expected, we observe clear spatial patterns (Figure 2). Overall, the projected death rates are much lower in the major UK cities and their surrounding areas, i.e. London, Bristol, Cardiff, Birmingham, Manchester, Liverpool, Glasgow and Edinburgh. These all are areas with relatively young population. In contrast, the projected mortality rates are high in small towns and rural areas. The settlement geography of Britain also leads to distinct regional patterns. Relatively high fatality rates are projected in large areas of South-West England (in Devon and Cornwall, also in Somerset and Dorset), in Central and North Wales and in Northern England (Cumbria, Northumbria and North Yorkshire). Interestingly, the expected death rates are also high in some coastal areas of East and South-East England (e.g. Norfolk and Sussex) showing the spatial clustering of old (and very old) population. In Scotland, fatality rates are high in Southern Scotland (Dumfries and Galloway, South Ayrshire and Scottish Borders) and in North-West Scotland (Highlands and Islands).

**Figure 2.**
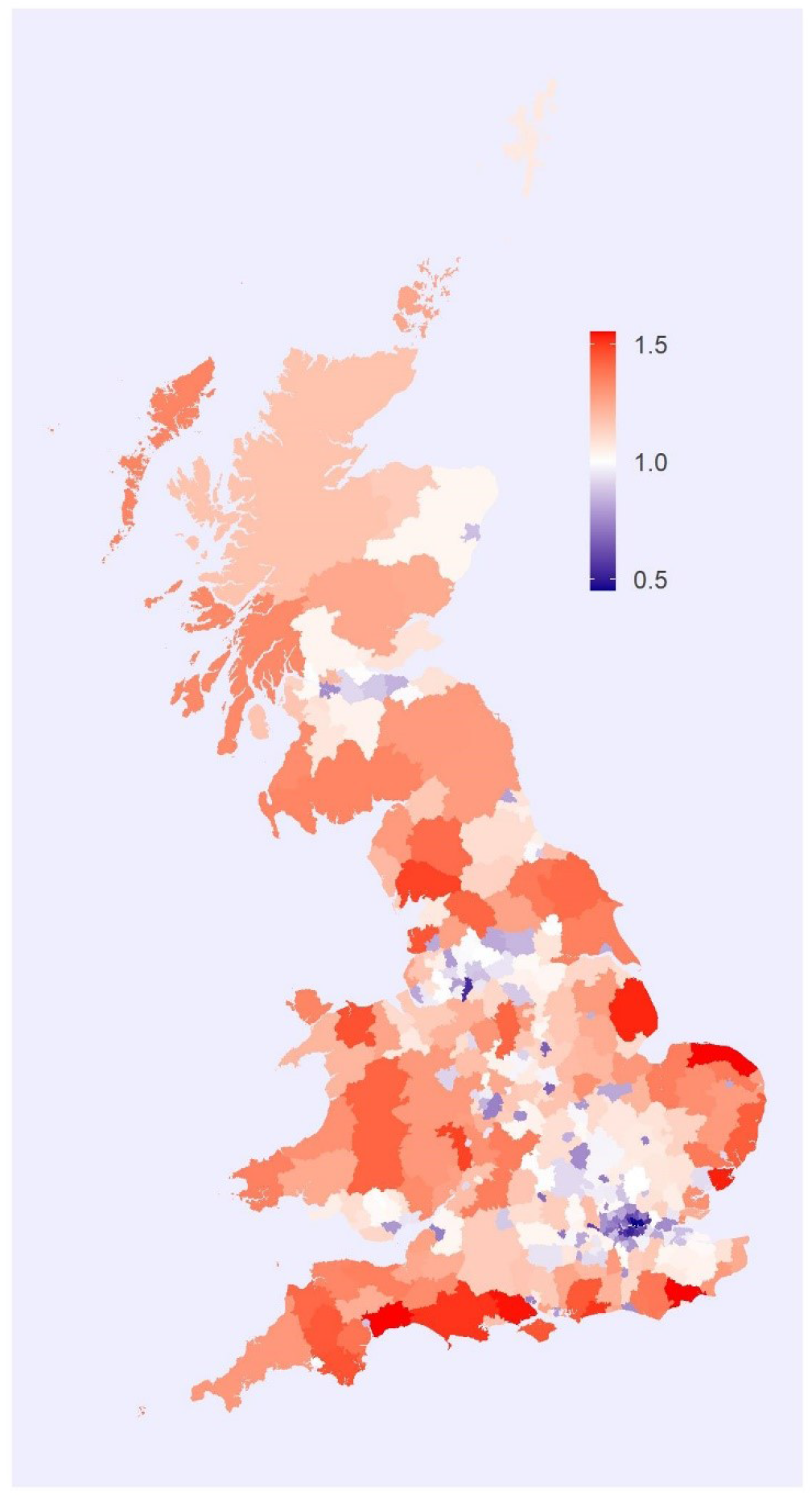
Projected relative infection fatality rates from Covid-19 in Great Britain by local authority districts.

### Small-area level

We have calculated the values of the two measures of spatial autocorrelation using the LSOA level data for England and Wales and Data Zones for Scotland. The value of Moran’s *I* is 0.58, which suggests a significant spatial autocorrelation in the projected infection fatality rates, i.e. areas with high and low fatality rates tend to cluster because of the high residential separation of different population age-groups in Great Britain. Figure 3 displays the clusters of relatively high (and also low) fatality rates based on the values of the local Moran’s *I* statistics. Clearly, we observe high fatality rates in a number of larger regions in England, Wales and Scotland we already discussed above; the analysis also reveals many ‘pockets’ with high fatality rates within the areas of relatively low death rates. (Figure A3 in Appendix shows the same calculations at the MSOA level with the value of Moran’s *I* of 0.65.) Figures 4a to 4d present the projected death rates for selected UK’s regions to illustrate variation within regions and the local clusters of high fatality rates. London has significantly lower projected death rates from Covid-19 than most other regions of the UK because of young population (Figure 4a). However, there are also areas with relatively high infection fatality rates in the London region, especially in several boroughs on the edge of the city region including Bromley and Croydon in the South, Havering in the East and parts of Hillingdon, Harrow and Barnet in the North. Figure 4b shows projected relative infection death rates in East of England. Again, we see significant variation within the region. There are pockets with expected high and low fatality rates in the Southwestern part of the region (e.g. Essex and Cambridgeshire), whereas the Eastern part of the region (counties of Norfolk and Suffolk), especially the seaside communities have relative high expected infection fatality rates, suggesting a high concentration of old population in these areas.

**Figure 3.**
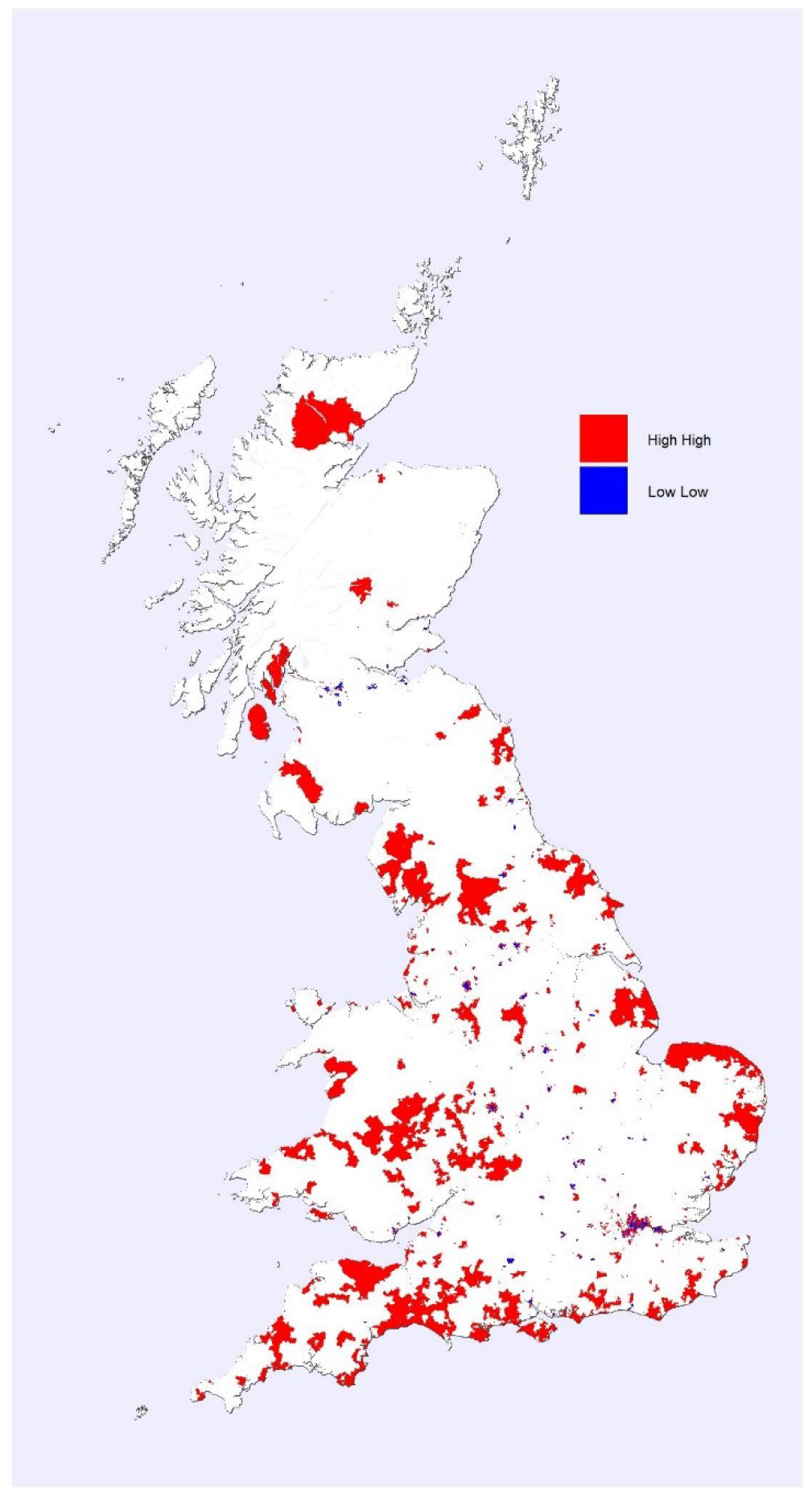
Projected clusters of high and low infection fatality rates from Covid-19 in Great Britain.

**Figure 4.**
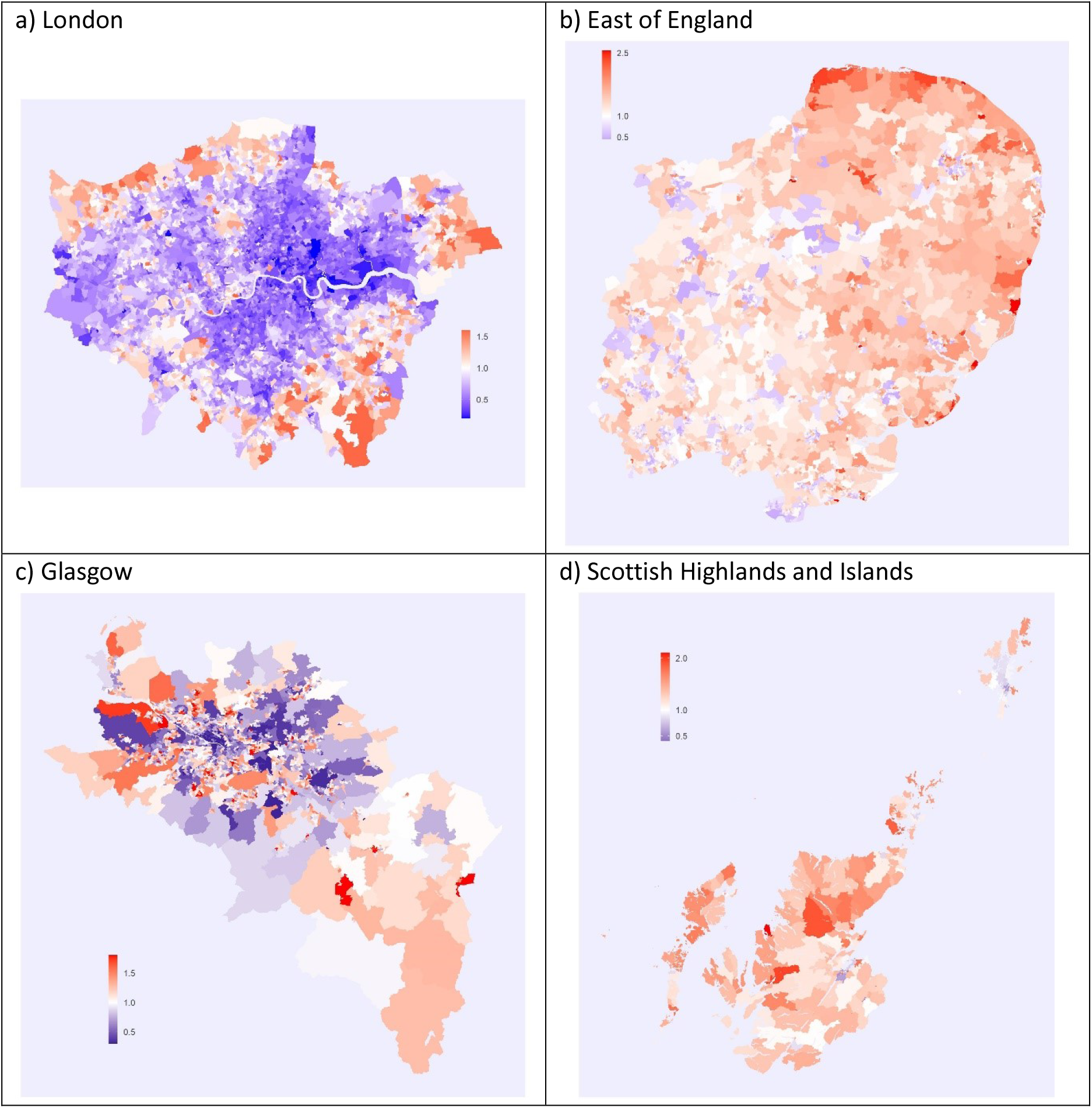
Projected relative infection fatality rates from Covid-19 in the UK for selected regions.

Similar spatial clustering and variation within regions is observed in Scotland. Projected infection fatality rates are lower than the UK’s average in Glasgow, the largest city of Scotland, but they vary within the city region (Figure 4c). The projected death rates are relatively low in the city centre and surronding areas with relatively young population and high in the Nortwestern and Southeastern part of the urban region. Again, there are many pockets with high fatality rates also in the city centre. The projected death rates from Covid-19 are relatively high in the Scottish Highlands (Figure 4d). Only the main urban area, i.e. the city of Inverness displays low levels. Interestingly, there is a large area with low expected fatality rates in Shetland, which extends from the main town of Lerwick to the north of the Mainland island.

Finally, we have calculated infection fatality rates by area type, separately for England and Wales and for Scotland, using the ONS and NRS urban-rural classifications of lower census output areas. Although the results by area type do not capture all regional variation in the projected infection fatality rates, they provide a good summary of the Britain’s settlement geography. The analysis shows that most deaths are expected to occur in urban areas; this is not surprising as the majority of the UK’s population lives in cities (Table 2). However, a different picture emerges when we calculate the number of expected deaths relative to the population size. The projected death rates from Covid-19 are significantly higher in small towns and rural settlements. The highest death rates are projected in remote small towns and rural areas; infection fatality rates there are as much as 85% and 55% higher than those in major urban areas in England and Scotland, correspondingly. In Scotland, these are communities with the driving time more than 30 minutes to a nearest settlement with 10 thousand or more inhabitants.

**Table 2.** Projected Proportion of Deaths and Relative Infection Fatality Rates from from Covid-19 in Britain by Area Type.

| Urban Rural Classification | Deaths | Fatality | Urban Rural Classification | Deaths | Fatality |
| --- | --- | --- | --- | --- | --- |
| *England and Wales* |  |  | *Scotland* |  |  |
| Urban major conurbation | 0.275 | 1 | Large urban areas | 0.30 | 1 |
| Urban minor conurbation | 0.032 | 1.15 | Other urban areas | 0.37 | 1.18 |
| Urban city and town | 0.457 | 1.27 | Accessible small towns | 0.09 | 1.27 |
| Urban city and town in a sparse setting | 0.003 | 1.62 | Remote small towns | 0.03 | 1.47 |
| Rural town and fringe | 0.116 | 1.57 | Very remote small towns | 0.01 | 1.43 |
| Rural town and fringe in a sparse setting | 0.008 | 1.84 | Accessible rural | 0.12 | 1.28 |
| Rural village and dispersed | 0.096 | 1.64 | Remote rural | 0.04 | 1.48 |
| Rural village and dispersed in a sparse setting | 0.014 | 1.85 | Very remote rural | 0.04 | 1.55 |

## Conclusions and discussion

The objective of this study was to investigate the contribution of population age structure to mortality from Covid-19 in the UK by geographical units. This is the first study to investigate the contribution of age structure to infection fatality rates within a country at different spatial scales. In many industrialised countries including the UK spatial segregation between different age groups is substantial. If we know how the infection fatality rates vary by age, we can project with a high accuracy expected death rates in geographical units.

Our analysis showed a significant variation in the projected death rates between the constituent countries of the UK, between its regions and within regions. First, Scotland and especially Wales exhibit higher projected fatality levels from Covid-19 than England, whereas Northern Ireland displays lower rate. These mortality differences reflect differences in the population age structure across the UK’s countries: Northern Ireland has the youngest age structure, whereas Scotland and Wales have the oldest. Second, the projected infection fatality rates are high in small towns and rural areas, which include e.g. large areas of South-West England, coastal areas of South-East England, Central and North Wales, Northern England, Southern Scotland and North-West Scotland. These all are regions with relatively old populations. In contrast, projected mortality rates are low in the major UK cities and their surrounding areas where young and middle-age population dominates. Overall, although the absolute number of deaths is still expected to be the largest in urban areas where most of the UK’s population lives, the death rates (or the number of expected deaths per 1,000 population) are projected to be 50-80% higher in remote small towns and rural areas than those in large urban areas. Third, our analysis showed that within urban areas there are ‘pockets’ with high projected death rates suggesting substantial residential segregation between the different age groups also within urban regions.

The study shows a significant geographical variation in projected death rates from Covid-19 in the UK and the clustering of areas with high and low infection fatality rates. Clearly, the results could be used by the NHS to plan resource allocation if the pandemic is to last long and the virus is to spread to all corners of the UK. Our study, which has identified the location of populations at high risk and identified vulnerable communities, shows that possibly there will be long-term socio-cultural impacts of the pandemic in addition to short-term public health and economic effects. If the virus is to spread widely to remote communities in the Scottish Highlands and Islands, Central and North Wales and Cornwall with relatively old populations, the Welsh-, Gaelic- and Cornish-speaking populations are projected to experience heavy population losses, which in the case of the two latter groups may challenge the very existence of these unique linguistic communities with long histories.

In our calculations we assumed that infection rate will be the same across regions. Previous research has shown that this is unlikely the case in the short-term; the spread of an infectious disease normally follows geographical patterns rather than is uniform across space. Our results thus show what would happen to different regions and communities if Covid-19 spread there. In the long-run the assumption of a uniform infection rate can be assumed for Covid-19 unless regional lockdowns are introduced, or the vaccine becomes rapidly available. We have also conducted series of robustness checks to test sensitivity of the results to different assumptions on age-specific fatality rates. Following research by Verity et al. we conducted projections with slightly higher and slightly lower IFRs rates (5). The results of these analyses were very similar to the main results supporting a significant variation in projected fatality rates by geographical units in the UK because of substantial residential age segregation.

National Statistical Agencies (ONS, NRS and NISRA) have rapidly responded to the pandemic by providing in their weekly mortality reports information on the location and age distribution of deaths by sex from Covid-19. We recommend that the age distribution of deaths by sex will also be provided for each local authority district or council area. Relevant public health agencies in the UK should follow the example and provide data on the number of reported infections and hospitalised people cross-classified by age, sex and area at minimum (e.g. clinical commissioning group or health board). Access to disaggregated data would allow scientific community to rapidly respond to the challenges by estimating the spread of the virus and by conducting data-based predictions on infected people, hospitalisations and deaths, which is much needed in the rapidly changing circumstances. The analysis of the weekly mortality statistics in London (collected by the parishes during the plague outbreaks in the late 16th century) by John Graunt and a mapping of the deaths by John Snow during a cholera epidemics in London in 1854 are excellent examples from the history how data, if made available, can be used by scientists for public good.

## Data Availability

All data-sets are available at the ONS and NRS websites.

## Contributions

HK and PD both conceptualised and designed the study. PD conducted data analysis and HK prepared a manuscript, which PD revised.

## Appendix

**Figure A1.**
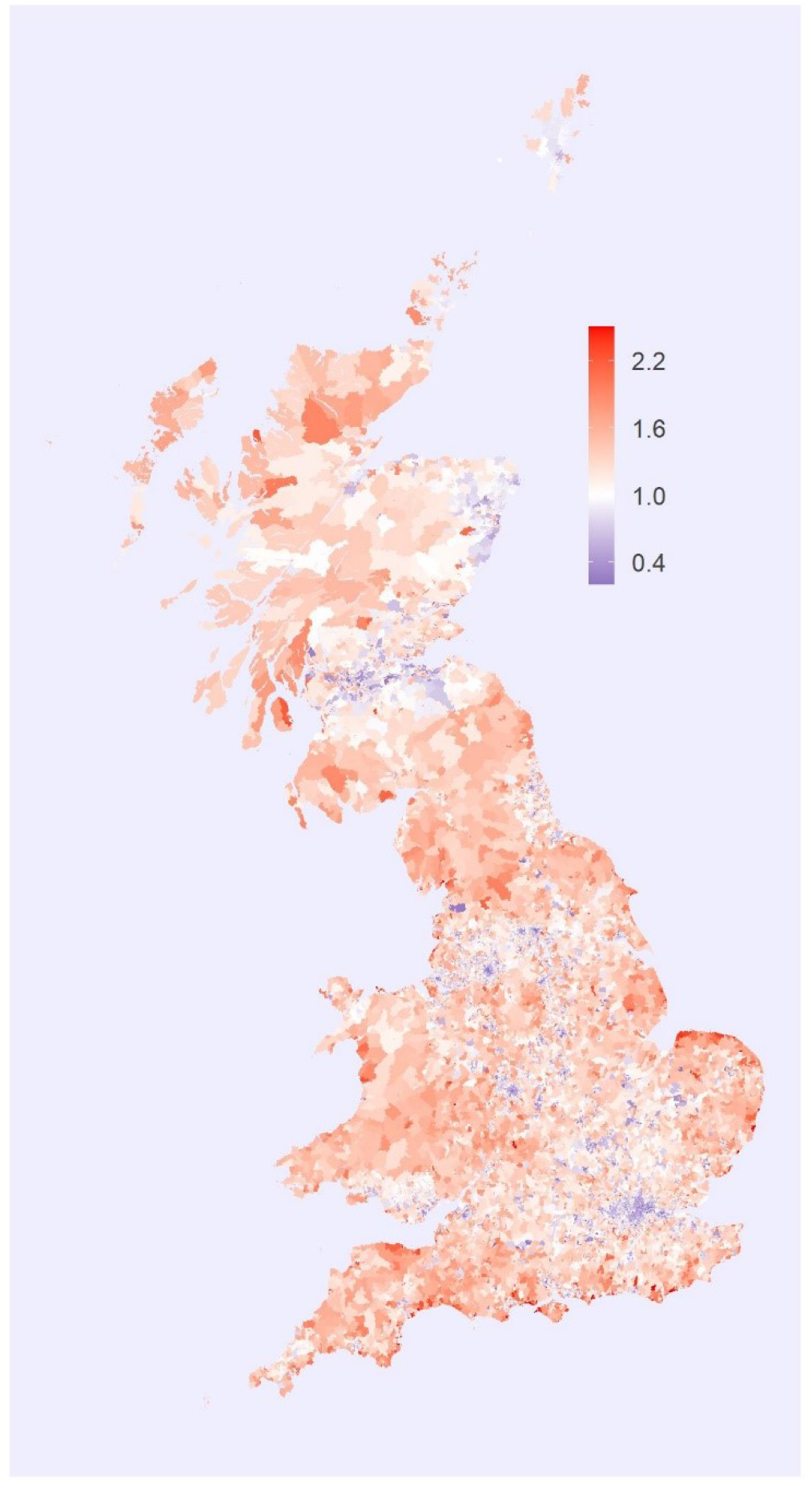
Projected relative infection fatality rates from Covid-19 in Great Britain by census lower super output areas.

**Figure A2.**
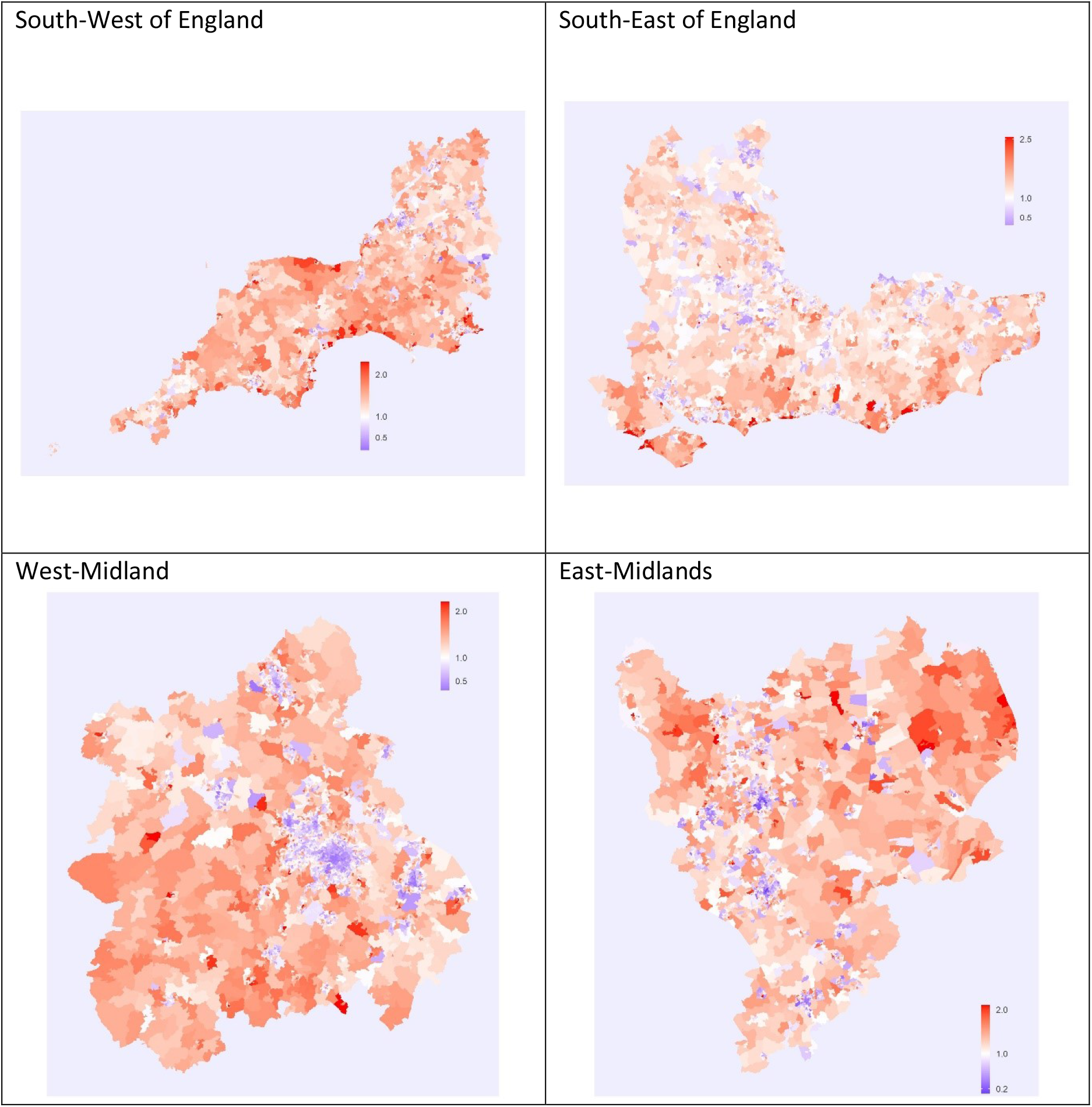
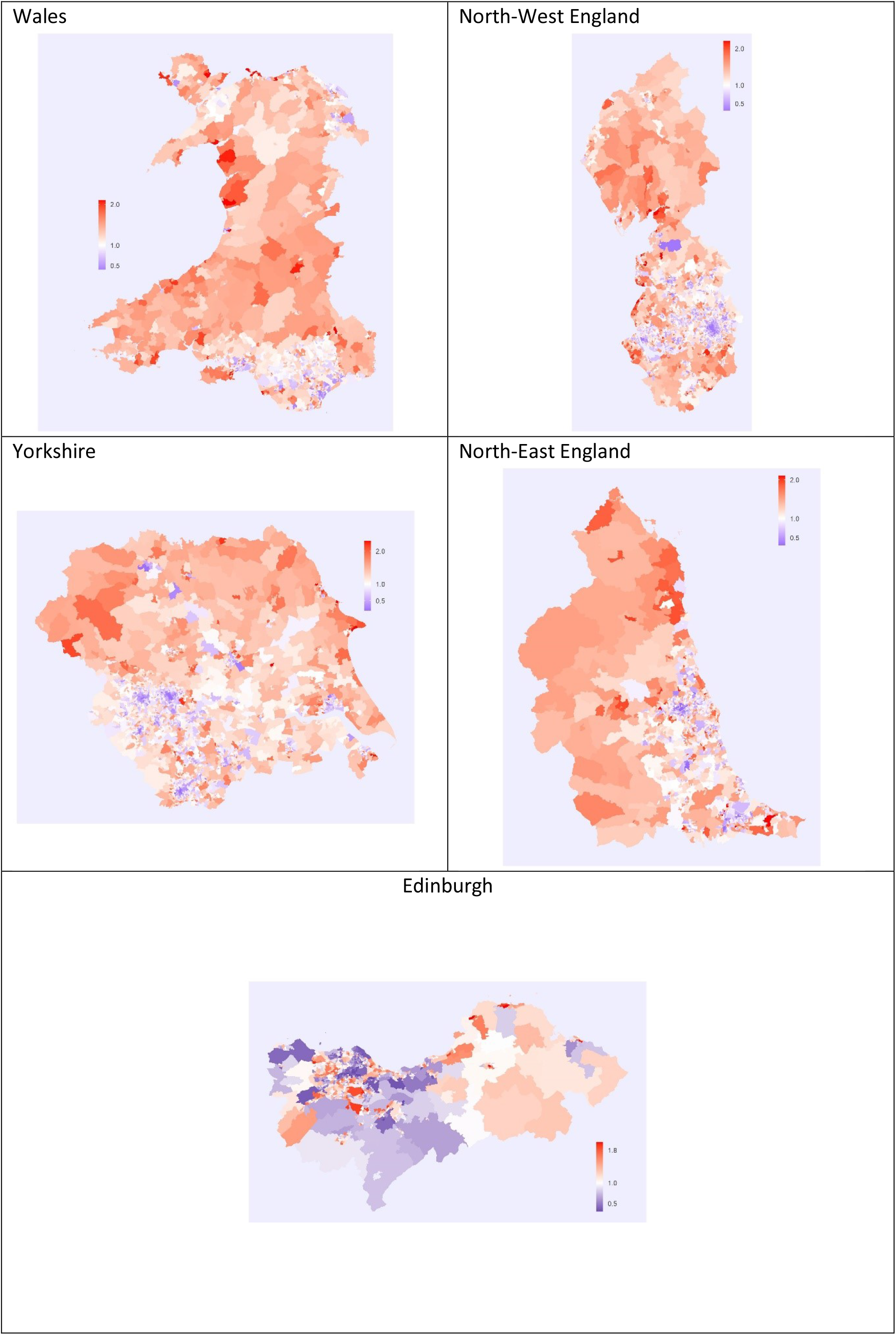
Projected relative infection fatality rates from Covid-19 in the UK for selected regions.

**Figure A3.**
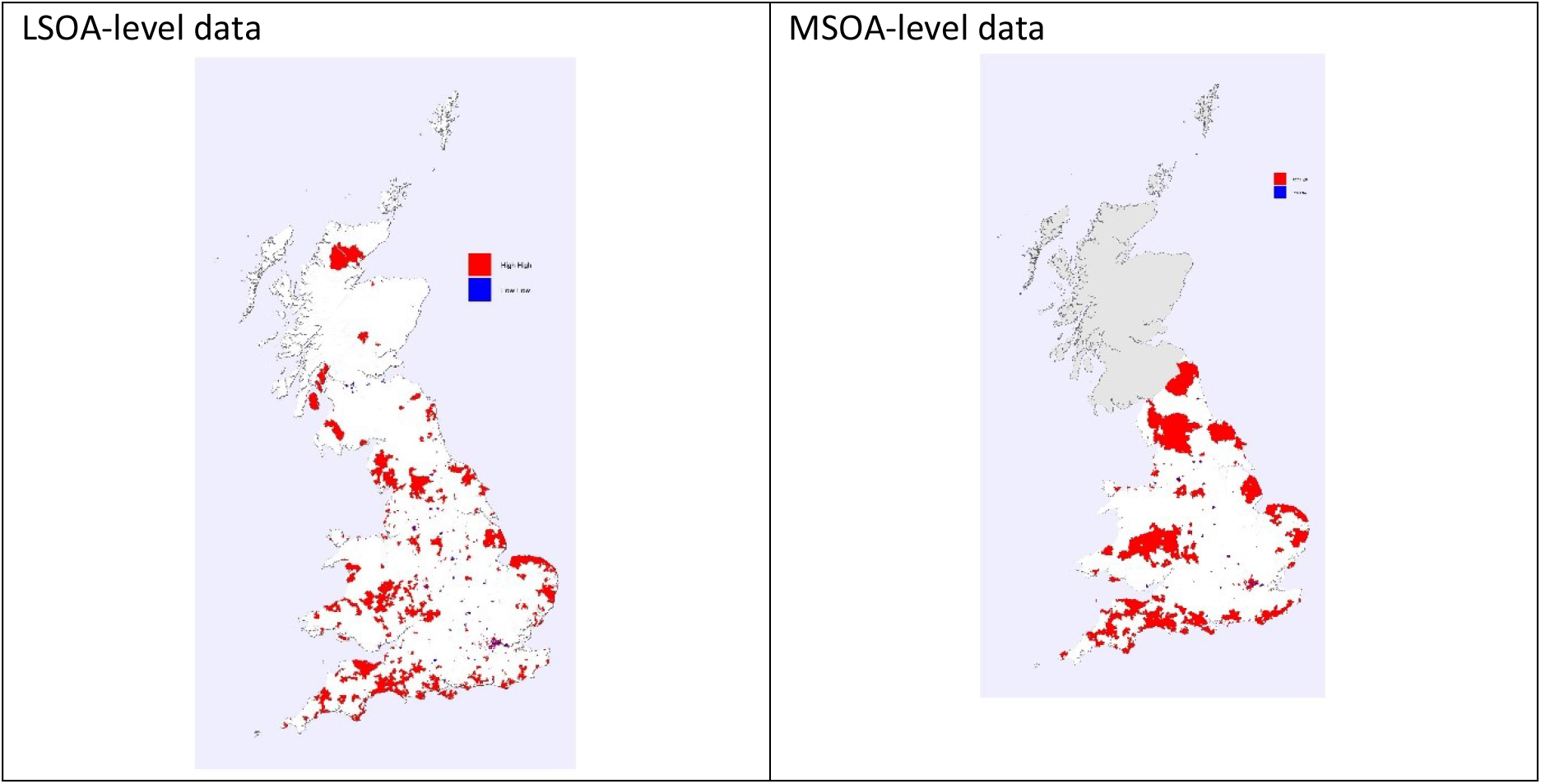
Comparison of projected clusters of high and low infection fatality rates from Covid-19 in Great Britain.

**Figure A4.**
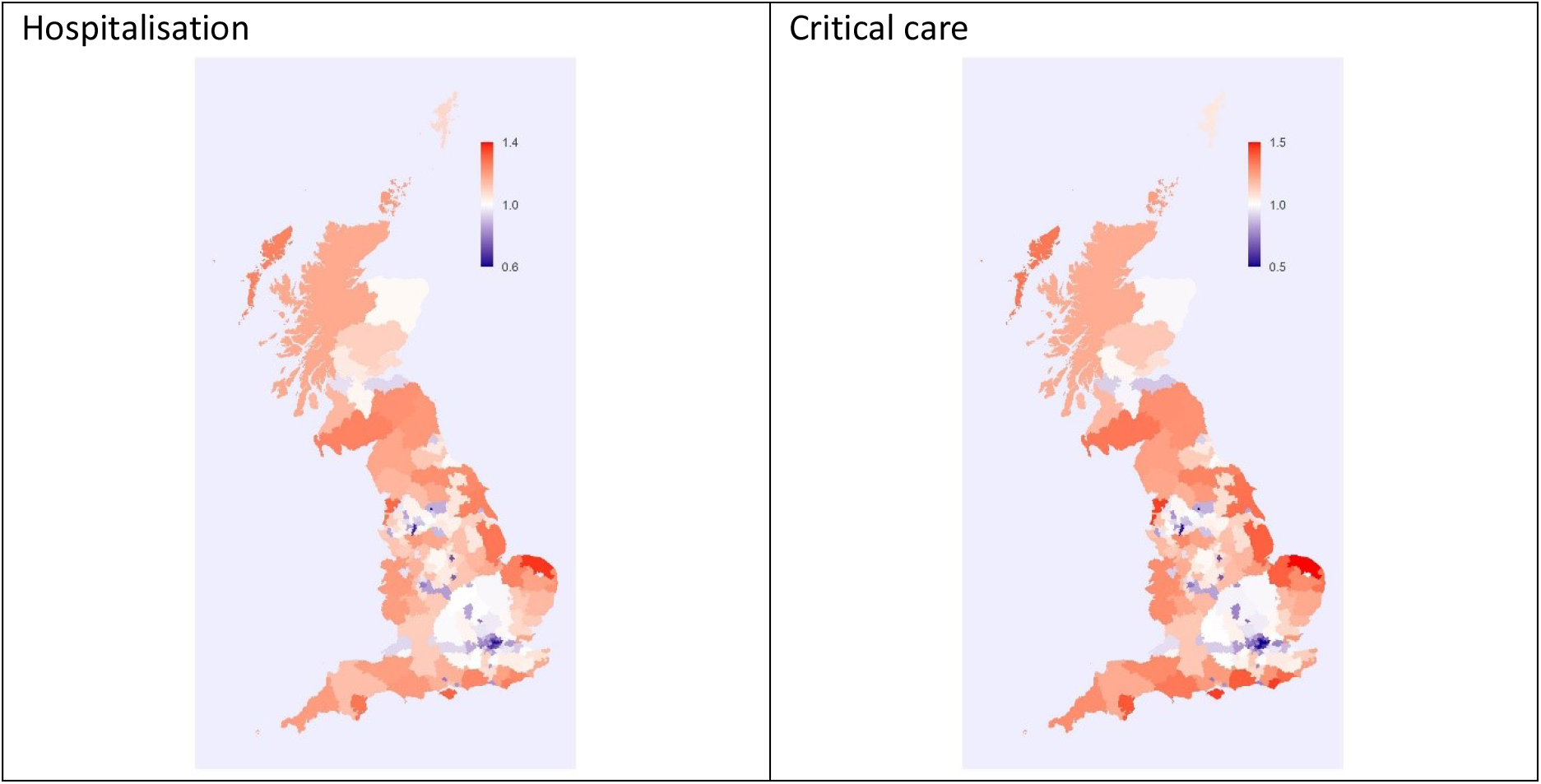
Projected relative hospitalisation and critical care rates from Covid-19 in England and Scotland by clinical commissioning groups and health boards.

## Footnotes

1 Great Britain experienced more than 4,400 registered deaths by 3/4/20 from Covid-19. The deaths of males formed between 57% (GB) and 61% (England and Wales) of the total number of deaths. We obtained the age-specific IFRs by sex by multiplying the age-specific IFRs by the coefficients of 1.2 and 0.8, respectively.

